# “Pandemic of the unvaccinated”? At midlife, white people are less vaccinated but still at less risk of Covid-19 mortality in Minnesota

**DOI:** 10.1101/2022.03.02.22271808

**Authors:** Elizabeth Wrigley-Field, Kaitlyn M. Berry, Andrew C. Stokes, Jonathon P. Leider

## Abstract

**Introduction:** Recent research underscores the exceptionally young age distribution of Covid-19 deaths in the United States compared with international peers. This brief characterizes how high levels of Covid mortality at midlife ages (45-64) are deeply intertwined with continuing racial inequity in Covid-19 mortality.

**Methods:** Mortality data from Minnesota in 2020-2022 were analyzed in June 2022. Death certificate data and published vaccination rates in Minnesota allow vaccination and mortality rates to be observed with greater age and temporal precision than national data.

**Results:** Black, Hispanic, and Asian adults under age 65 were all more highly vaccinated than white populations of the same ages during most of Minnesota’s substantial and sustained Delta surge and all of the subsequent Omicron surge. However, white mortality rates were lower than those of all other groups. These disparities were extreme; at midlife ages (ages 45-64), during the Omicron period, more highly-vaccinated populations had COVID-19 mortality that was 164% (Asian-American), 115% (Hispanic), or 208% (Black) of white Covid-19 mortality at these ages. In Black, Indigenous, and People of Color (BIPOC) populations as a whole, Covid-19 mortality at ages 55-64 was greater than white mortality at 10 years older.

**Conclusions:** This discrepancy between vaccination and mortality patterning by race/ethnicity suggests that, if the current period is a “pandemic of the unvaccinated,” it also remains a “pandemic of the disadvantaged” in ways that can decouple from vaccination rates. This result implies an urgent need to center health equity in the development of Covid-19 policy measures.

## Introduction

The introduction of effective vaccines for Covid-19 in early 2021 resulted in a general consensus, for a time, that future pandemic surges would lead to a “pandemic of the unvaccinated.”^1^ However, limited data are available to evaluate this characterization for different racial/ethnic groups, despite substantial evidence that vaccination status and mortality both differ dramatically by race/ethnicity across the US.^2,3^ In the present study, we evaluated whether Covid-19 mortality is reflective of vaccination rates for different racial/ethnic groups in Minnesota using death certificate data on all Covid-19 deaths from March 2020 to April 2022. Recent research emphasizes the exceptionally young age distribution of Covid-19 deaths in the United States relative to other countries.^4,5^ Because deaths at midlife ages drove this phenomenon and because such deaths exhibited substantial racial/ethnic inequality before vaccines were available,^6^ we focus on vaccination and mortality at these key ages.

## Methods

This analysis uses death certificate data from Minnesota, March 2020 through April 2022; state vaccination data; and National Center for Health Statistics population distributions. We examined Minnesota because of the unique availability of near-real-time data on both vaccination status and Covid-19 mortality that are simultaneously separated by race/ethnicity and age. In contrast, national vaccination data that are race/ethnicity-specific are not separated by age. Minnesota also stands out for its prolonged and deadly surge of the Delta variant, which did not end until it was supplanted by the Omicron surge at the end of 2021.^7^ We examined mortality patterns in specific racial/ethnic groups at midlife, with a particular focus on periods of high mortality following widespread vaccination in Fall 2021 and Spring 2022. Covid-19 mortality patterns in Minnesota justify our analytic grouping of the state’s Black, Indigenous, and People of Color (BIPOC) population, as elaborated in the Appendix.

Deaths were defined as Covid-19 deaths if there was any mention of U07.1 on the death certificate. We calculated Covid-19 death and vaccination rates by race/ethnicity and age for four pandemic periods corresponding to pre-vaccination (March 2020-January 2021); mid-vaccination (February-June 2021); Delta-dominated (July-December 2021); and Omicron-dominated (January-April 2022) periods. We used “fully vaccinated” rather than “single-shot vaccinated” as our vaccination metric because full vaccination provides greater protection and is likely reported with higher data quality.^8^ Booster shot uptake is not available by race/ethnicity in Minnesota. BIPOC vaccination at elderly ages is likely underestimated in these data, as discussed in the Appendix, which outlines additional methodological considerations.

## Results

By the end of 2021 in Minnesota, vaccination among white Minnesotans was outpaced by vaccination among Black, Indigenous, and People of Color (BIPOC) Minnesotans at midlife ages (45-64) as well as young adult ages (19-44). Yet in all age groups and in each phase of the pandemic, white mortality was substantially lower than mortality among Minnesotans of color (Figure 1).

**Figure 1.**
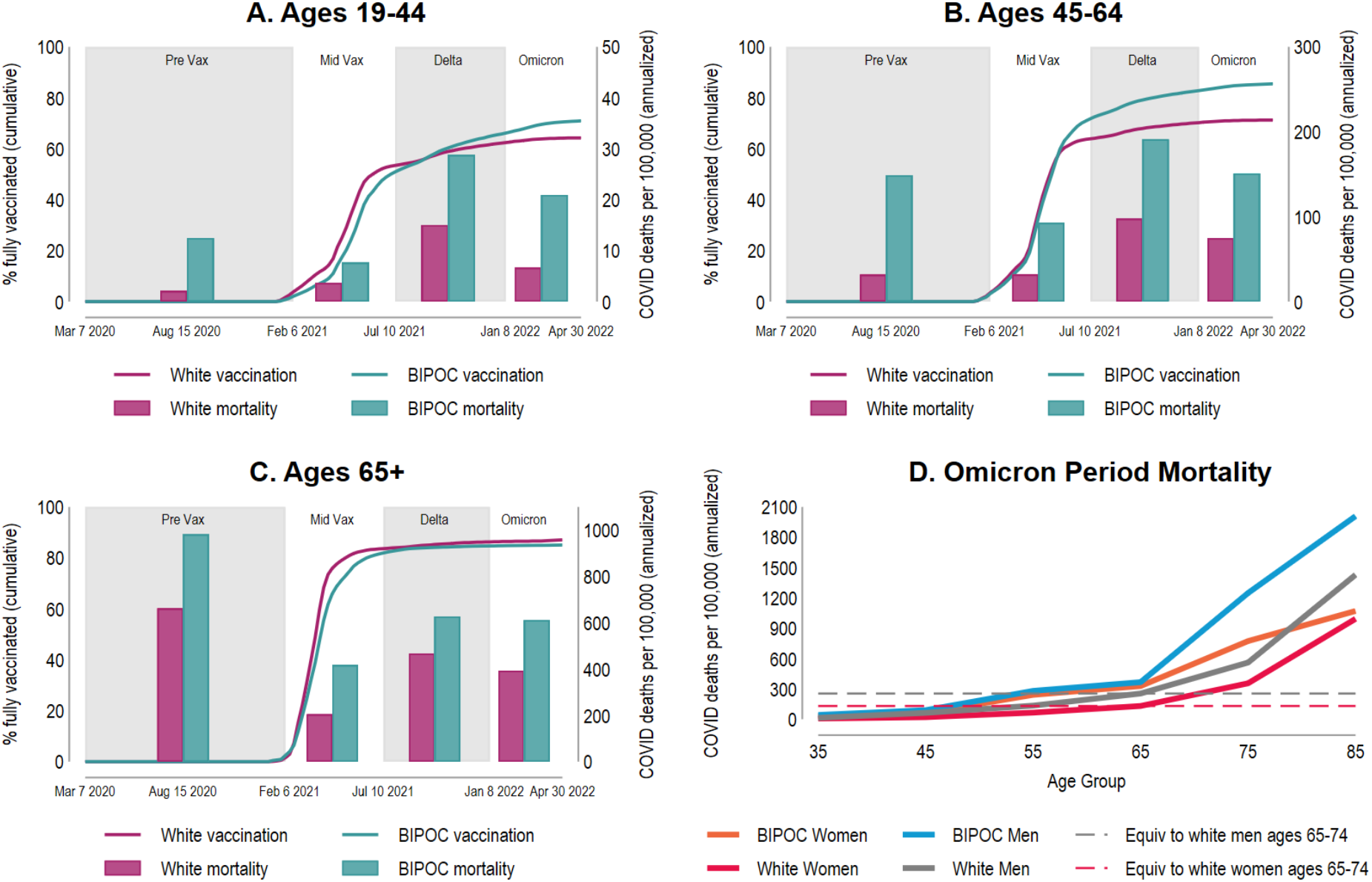
Period-specific vaccination rates and Covid-19 mortality rates for white and BIPOC Minnesotans by age. The lines in panels A-C depict the cumulative vaccination progress of each group over time as measured by the percentage of that race-specific, age-specific group that have completed their vaccine series (left axis). The bars in panels A-C depict the race-specific, age-specific mortality rates (right axis) during the pre-vaccination period, mid-vaccination period, Delta period and Omicron period of the pandemic. Mortality rates are annualized to facilitate comparison across periods. Vaccination rates and mortality rates are presented for Minnesotans ages 19-44 (Panel A), ages 45-64 (Panel B), and ages 65+ (Panel C). Panel D shows how, during the Omicron period, Covid-19 mortality rates increase with increasing age for BIPOC women, white women, BIPOC men, and white men. The dashed lines show the relatively lower age at which BIPOC groups experience the same mortality rates as white groups at ages 65-74.

White undervaccination at midlife ages is pronounced: at the end of April 2022, “fully vaccinated” rates were 85% for BIPOC Minnesotans compared to only 71% for white Minnesotans (Figure 1B). Midlife vaccination for BIPOC Minnesotans is similar to vaccination rates for elderly (65+) white Minnesotans (87%; Figure 1C). Yet, the gap in BIPOC-white mortality at those midlife ages was extreme; for example, during the Delta and Omicron periods, BIPOC mortality at ages 55-64 was higher than white mortality at ages 65-74 (Figure 1D, Figure S3, Appendix Table S1). At key midlife ages, Minnesota’s white population is its second-least vaccinated racial/ethnic group, following Native Americans (Figure 2). However, despite low vaccination rates, Minnesota’s white population aged 45-64 has lower mortality than that of all other racial/ethnic groups, which ranged, during the period dominated by the Omicron variant, from 115% (Hispanic) to 661% (Native) of white mortality (Appendix Table S2).

**Figure 2.**
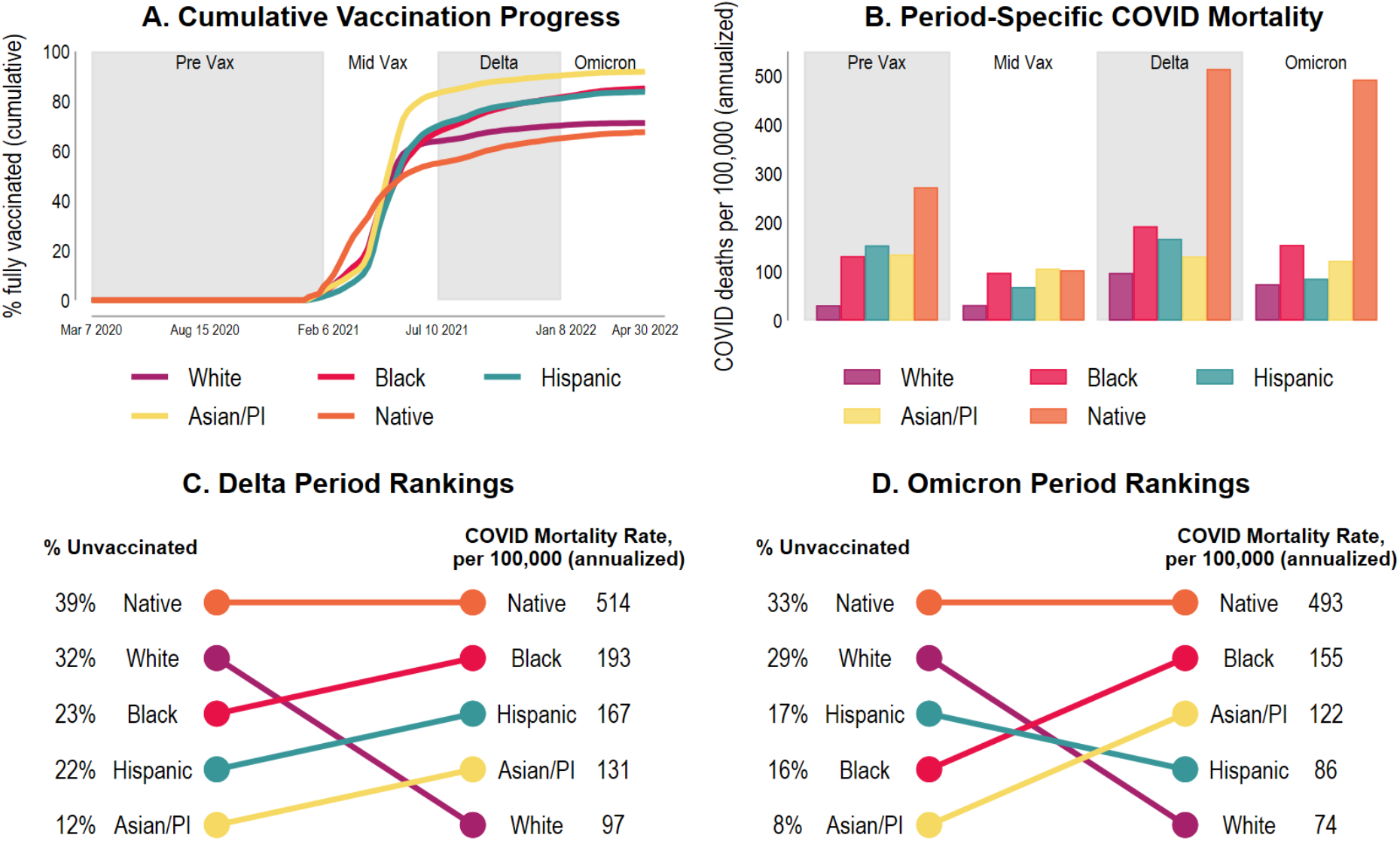
Period-specific vaccination rates and Covid-19 mortality rates at midlife (ages 45-64) by race/ethnicity in Minnesota. Panel A shows the cumulative vaccination progress of each racial group over time at midlife as measured by the percentage of that group who have completed their vaccine series. Panel B depicts the race-specific mortality rates at midlife during the pre-vaccination period, mid-vaccination period, Delta period, and Omicron period of the pandemic. Panel C-D compares the ranking of racial groups, from worst to best performing, by percent unvaccinated and mortality rates during the Delta period (Panel C) and Omicron period (Panel D). The vaccination data in Panels C-D is from the midpoint of each period (October 2, 2021 for Delta; February 26, 2022 for Omicron) and the mortality rates cover the entire period (July 2021 to December 2021 for Delta; January 2022 to April 2022 for Omicron). All mortality rates are annualized to facilitate comparison across periods.

## Discussion

The present study found that despite lower vaccination rates than all but Native Minnesotans, white Minnesotans had lower Covid-19 mortality at midlife than did Black, Hispanic, Asian, and Native Minnesotans from autumn 2021 through April 2022. We note two broad possible explanations for these results. One possibility is that racial inequity in Covid-19 mortality risk—due to differential transmission, comorbidities, or unequal medical access^9^— among the unvaccinated, the vaccinated, or both, may be so great that it overwhelms the differences in vaccination status. A second possibility is that findings may reflect vaccine differences within the “fully vaccinated” population, with people of color potentially less likely to have received booster shots and less likely to have received mRNA vaccines in their primary series.^10^

Regardless of the precise mechanism, the findings suggest that the “pandemic of the unvaccinated” formulation is incomplete and that Covid-19 also remains a “pandemic of the disadvantaged.” If population mortality primarily reflected population vaccination rates, white communities in midlife would have a greater burden of Covid-19 mortality than communities of color. The fact that we observed the opposite indicates that structural racism, as manifested through systems and policies that affect health care access, occupational risk, and housing conditions, continue to fundamentally shape risk of Covid-19 mortality even in the Delta/Omicron period.^11–14^

While a “pandemic of the unvaccinated” framing may be used as a rationale for accelerating a “return to normal,” a “pandemic of the disadvantaged” framing emphasizes the need for sustained population-based Covid-19 prevention and mitigation strategies that center health equity. Such measures could aim to further increase vaccination with community campaigns,^15^ and might also aim to mitigate Covid-19 spread through approaches that protect the vaccinated and unvaccinated alike, including improved ventilation in workplaces and public buildings, paid sick leave to prevent work while ill, Medicaid expansion and universal health care, economic payments to marginalized and medically high-risk populations, protective equipment and increased pay for long-term care workers so they have less need to work multiple jobs, eviction moratoriums and other housing support, continued mask mandates, and public funding for community testing programs and scientific research on Covid. These strategies acknowledge that, even when vaccine uptake among people of color is relatively high, the mortality of the pandemic remains unequally borne. The “pandemic of the disadvantaged” framing suggests that a sole emphasis on individual behavior is inadequate for reducing health inequities.

The extent to which findings in Minnesota may resemble those of other state contexts is unclear. If vaccination rates are generally higher in metropolitan areas compared with rural areas, other states with very urban populations of color and large rural white populations may show similar vaccination disparities. At the national level, aggregated over all ages, the white population is vaccinated at lower rates than all but African-American individuals,^16^ and in most states, white vaccination is lower than the high average age of white populations would predict.^17^ However, the lack of publicly-available data on the age composition of vaccine status by race/ethnicity for the U.S. as a whole limits the ability to know how widespread the midlife patterns identified here may be.

## Conclusions

The results in the present study highlight how this distinctive risk at midlife may be intertwined with the deep inequality in U.S. Covid-19 mortality. Populations of color may be at notably high risk at midlife—even when they have greater vaccination rates than white people at the same ages.

## Data Availability

All data produced are available online at https://osf.io/bxjkh/?view_only=80e912ce2b624ab7a78e51754c3952e3

## Acknowledgements

This research was supported by the Eunice Kennedy Shriver National Institute on Child Health and Human Development (P2CHD041023, F31HD107980), the National Institute on Aging (P30AG066613, R01AG060115-04S1), the Robert Wood Johnson Foundation (Grant #77521), and the University of Minnesota School of Public Health. We thank the Minnesota Department of Health, and particularly Keeley Morris, for sharing data and code that facilitated our analysis. The interpretations, conclusions, and recommendations in this work are those of the authors and do not necessarily represent the views of the National Institutes of Health, the Robert Wood Johnson Foundation, or the Minnesota Department of Health. We thank Michelle Niemann and Matthew Plummer for helpful comments.

## Data Availability

Data are available at https://osf.io/bxjkh/?view_only=80e912ce2b624ab7a78e51754c3952e3

## Methods Appendix

The death certificates analyzed were those included in a data release from the Minnesota Department of Health (MDH) on June 1, 2022. Among 2021 Covid-19 deaths included in the death certificate database, 91% were added to these regularly-updated data within three weeks following the date of death. Thus, these data are likely close to complete for included 2022 deaths (deaths occurring through April 30, 2022).

To create death rates from the death certificates, we used 2020 population estimates from the National Center for Health Statistics (NCHS), as the most recently available estimates. We chose not to project denominators for 2021, instead using official 2020 estimates, since 2020 population dynamics (shaping 2021 populations) are so distinctive relative to the dynamics of prior years. In order to report death rates across periods of unequal length (e.g., an 11-month pre-vaccination period vs. a 6-month Delta period), we annualized all rates. To construct age- and sex-standardized death rates at midlife for Figure S1A-B, we performed a “direct” standardization that reweights each race/ethnic population’s mortality using a shared set of weights.^18^ As our “standard” population weights, we used the 5-year-age and sex distribution of the full 2020 Minnesota population at ages 45-64.

Vaccine data were reported by MDH at the adult age categories 19-44, 45-64, and 65+; our main mortality analyses accordingly used these groups. Some racial group populations are too small to examine separately for ages 65+ in Minnesota. Thus, we show individual racial groups at ages 45-64 (Figure 2) and dichotomous white vs. BIPOC when examining all age groups separately (Figure 1). Mortality analyses support the use of the BIPOC category because, in Minnesota, the primary mortality divide at all high-mortality ages, all time periods, and for both women and men is between white Minnesotans vs. all others.^19^ In particular, Minnesota’s Asian/Asian-American population, the plurality of which is Hmong, is more disadvantaged in mortality than the Asian/Asian-American populations of states such as California.^6^ We report mortality rates using imputed race for multiracial populations; race is imputed using trumping rules in the order Hispanic ethnicity / Black / Asian / Native / Other / White. Thus, for example, someone whose death certificate reported Asian and White race would be recoded as Asian; someone reporting Black and Native race and Hispanic ethnicity would be recoded as Hispanic. In practice, these racial/ethnic assignments closely match NCHS race-bridged assignments (available through 2021 but not for 2022 deaths), as shown in Figure S2. In contrast, vaccination rates are reported by the state with single-race populations. We use denominators constructed with the appropriate racial categories for these respective numerators.

Minnesota vaccination data are reported as population percentages using population denominators estimated by the Minnesota Department of Health (MDH). These MDH denominators are derived from the 2019 American Community Survey (ACS) 5-year estimates (reflecting populations in 2015-2019) and Census Population Estimates Program (PEP) estimates. Because these denominators are several years old, they can result in unrealistically high vaccination estimates for growing subpopulations; for example, state data report that 100% of Black and Asian Minnesotans aged 65+ were fully vaccinated by August 2021. We adjusted these vaccine rates using the ratio of 2020 population sizes (as estimated by the NCHS) to the MDH estimates. We note that, because state vaccination data for Black and Asian populations aged 65+ are top-coded at 100%, our adjusted estimates likely under-estimate vaccination for these populations; moreover, under-estimating these two groups will produce likely under-estimation in the BIPOC category as a whole since Black and Asian populations together constitute 69% of the 65+ BIPOC population in Minnesota. This under-estimation is conservative for our argument because it implies that the seeming similarity in vaccination among white and BIPOC populations aged 65+ (Figure 1C) may disguise greater true vaccination among BIPOC populations, as among adults younger than age 65.

Sex-stratified results emphasize the extent of high COVID-19 mortality among Minnesota’s BIPOC population at midlife ages. During the Delta period for both men and women, and during the Omicron period for women, BIPOC mortality at ages 55-64 was higher than white mortality at ages 65-74, as was the case for white and BIPOC populations aggregated over sex (Table S1; compare with Figure 1D, Figure S3).

Due to the relatively old age of the white population even within the midlife age group, in age- and sex-standardized rates, the disparities in this period are even starker, with other racial groups ranging from 138% (Hispanic, Omicron) to 683% (Native, Omicron) of white mortality (Figure S2A-B).

## SUPPLEMENTAL TABLES

**Table S1.**
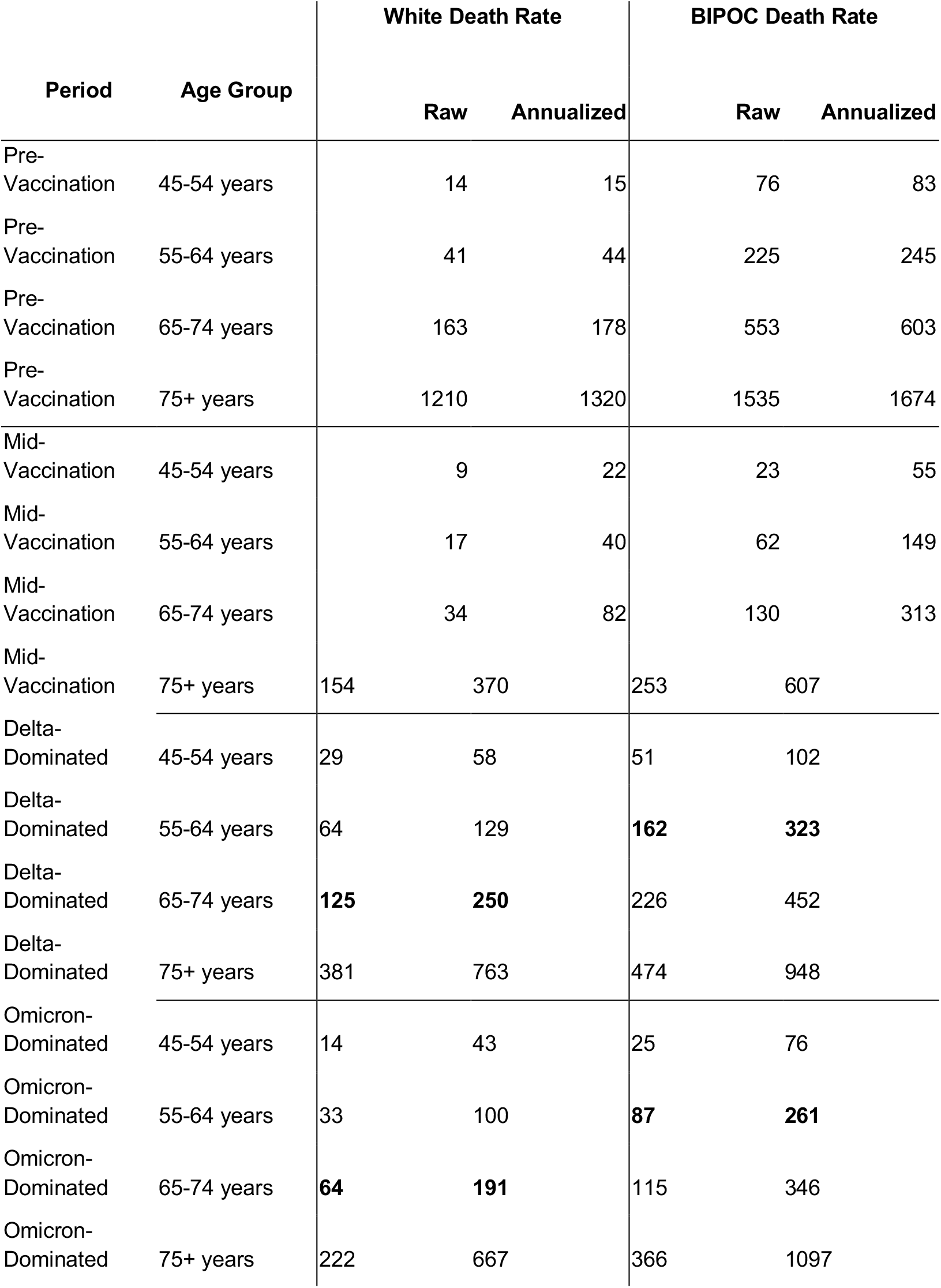
Covid-19 mortality by period, age, and race/ethnicity among midlife and elderly populations. Deaths are per 100,000 population. Bold numbers highlight the comparison made in the text between older-midlife BIPOC and younger-elderly white mortality rates.

**Table S2.** Mid-life (ages 45-64) vaccination and mortality by race and ethnicity.

| Period | Race/Ethnicity | Percent Unvaccinated<br>at Period Midpoint | Covid-19 Deaths<br>per 100,000 | Ratio to White<br>Mortality |
| --- | --- | --- | --- | --- |
| Pre-Vaccination | White | 100.00 | 31.39 |  |
| Pre-Vaccination | Black | 100.00 | 131.75 | 4.20 |
| Pre-Vaccination | Hispanic | 100.00 | 153.45 | 4.89 |
| Pre-Vaccination | Asian/PI | 100.00 | 135.05 | 4.30 |
| Pre-Vaccination | Native | 100.00 | 272.63 | 8.68 |
| Mid-Vaccination | White | 64.94 | 31.77 |  |
| Mid-Vaccination | Black | 67.65 | 97.70 | 3.08 |
| Mid-Vaccination | Hispanic | 71.47 | 68.43 | 2.15 |
| Mid-Vaccination | Asian/PI | 64.95 | 106.11 | 3.34 |
| Mid-Vaccination | Native | 59.59 | 102.82 | 3.24 |
| Delta-Dominated | White | 32.06 | 97.35 |  |
| Delta-Dominated | Black | 23.20 | 192.69 | 1.98 |
| Delta-Dominated | Hispanic | 22.26 | 167.28 | 1.72 |
| Delta-Dominated | Asian/PI | 12.03 | 130.87 | 1.34 |
| Delta-Dominated | Native | 39.03 | 514.10 | 5.28 |
| Omicron-Dominated | White | 28.95 | 74.49 |  |
| Omicron-Dominated | Black | 15.92 | 154.70 | 2.08 |
| Omicron-Dominated | Hispanic | 16.83 | 85.54 | 1.15 |
| Omicron-Dominated | Asian/PI | 8.50 | 122.03 | 1.64 |
| Omicron-Dominated | Native | 33.24 | 492.68 | 6.61 |

## SUPPLEMENTAL FIGURES

**Figure S1.**
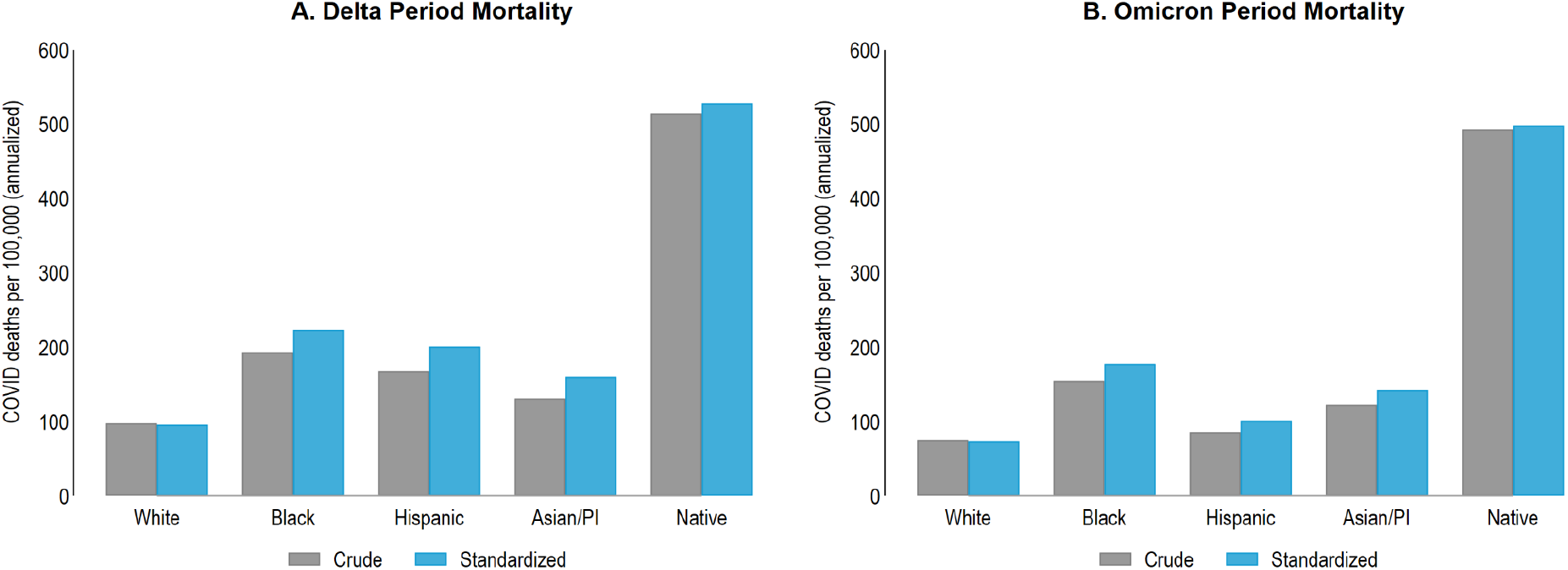
Delta-specific and Omicron-specific Covid-19 mortality rates and vaccination rates at midlife (ages 45-64) by race/ethnicity in Minnesota, with and without age and sex standardization. Panels A-B compare crude and age- and sex-standardized Covid mortality rates at midlife (ages 45-64) during the Delta period (Panel A) and the Omicron Period (Panel B).

**Figure S2.**
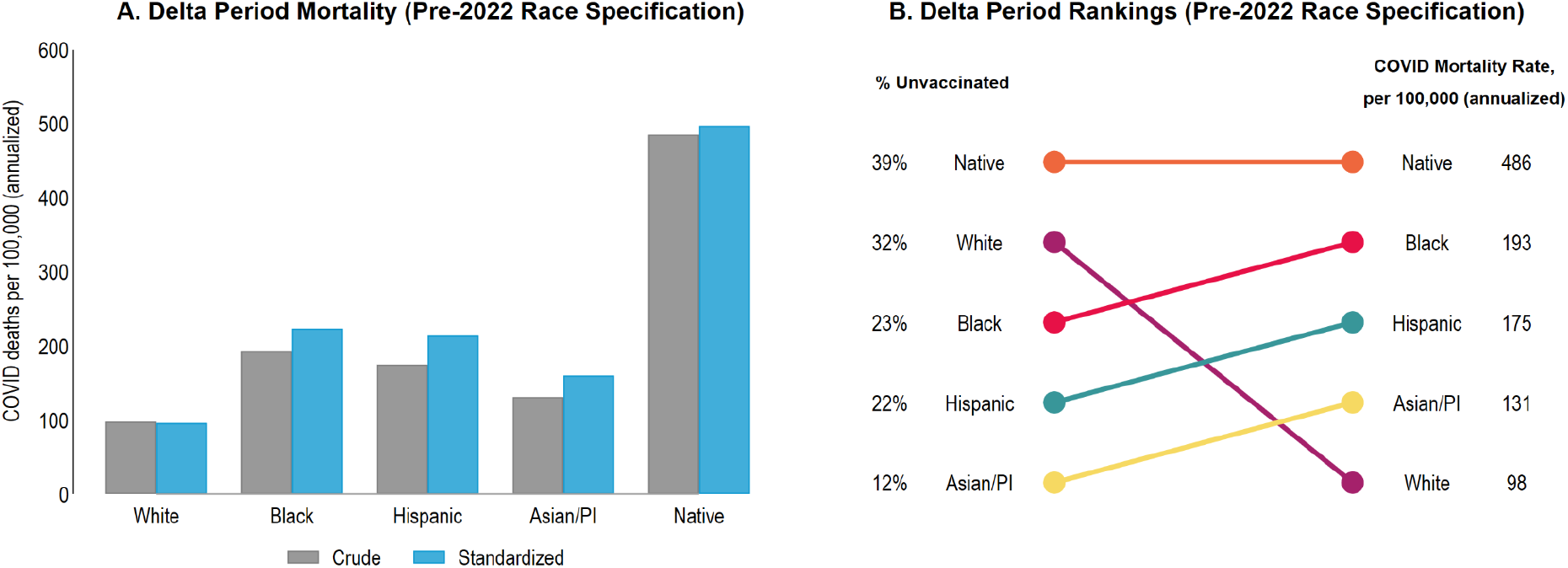
Delta-specific Covid-19 mortality at midlife (ages 45-64) with alternative racial/ethnic category assignments. Both panels use race-bridging racial/ethnic category assignments from the National Center for Health Statistics, which are unavailable for 2022. Panel A compares crude and age- and sex-standardized Covid mortality rates at midlife (ages 45-64) during the Delta period, and can be compared with Figure 2A. Panel B compares the ranking of racial groups, from worst to best performing, by percent unvaccinated and mortality rates, and can be compared with Figure 2C.

**Figure S3.**
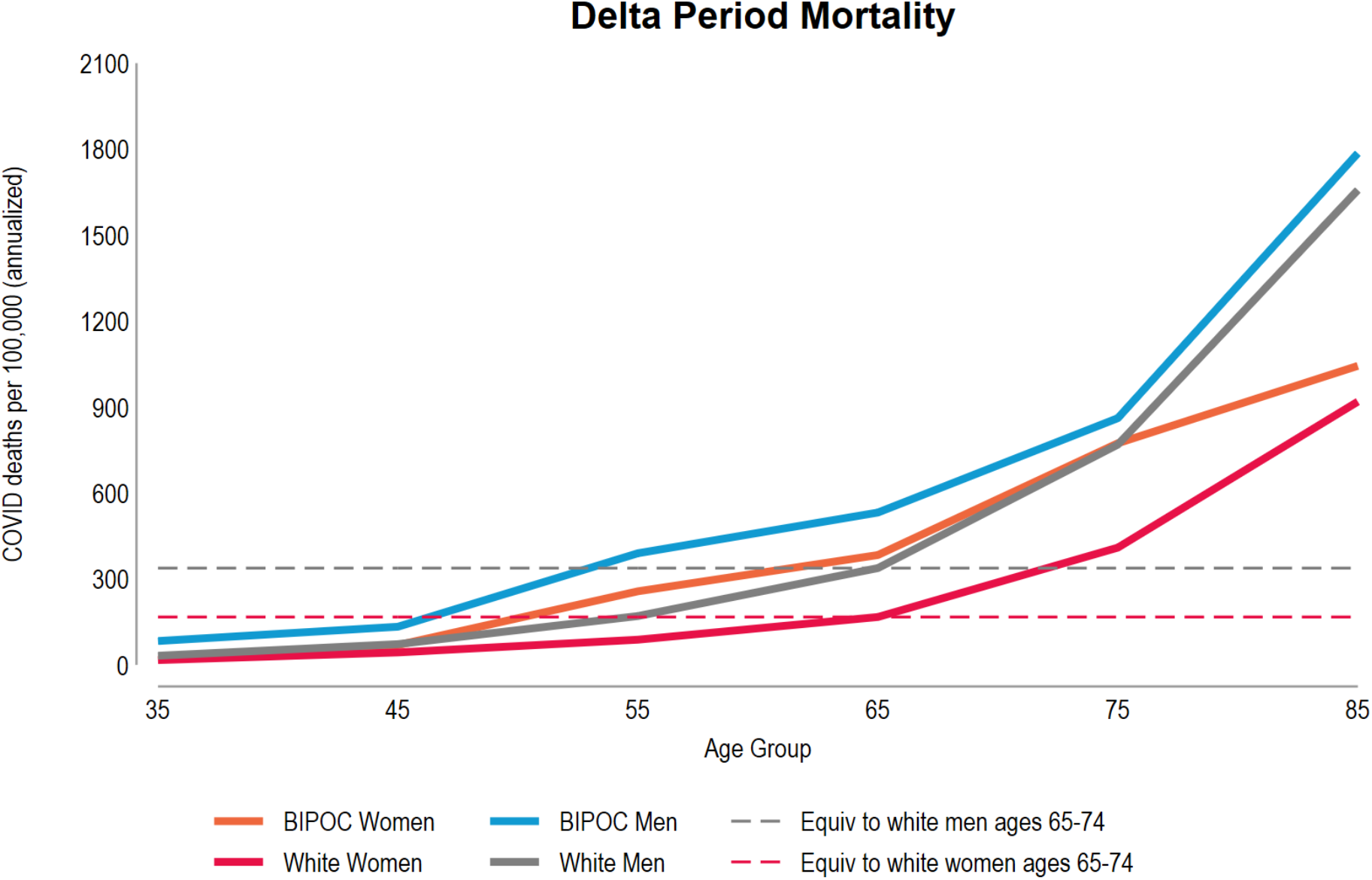
Mortality rates by age, race/ethnicity, and sex during the Delta period. Panel D shows how, during the Omicron period, Covid-19 mortality rates increase with increasing age for BIPOC women, white women, BIPOC men, and white men. The dashed lines show the relatively lower age at which BIPOC groups experience the same mortality rates as white groups at ages 65-74. This figure is the Delta-period version of Figure 1D.

## References

1. Sullivan B. U.S. COVID Deaths Are Rising Again. Experts Call It A “Pandemic Of The Unvaccinated.” NPR. https://www.npr.org/2021/07/16/1017002907/u-s-covid-deaths-are-rising-again-experts-call-it-a-pandemic-of-the-unvaccinated. Published July 16, 2021. Accessed February 18, 2022.

2. Ndugga N, Hill L, Artiga S, Haldar S. Latest Data on COVID-19 Vaccinations by Race/Ethnicity. Kaiser Family Foundation. https://www.kff.org/coronavirus-covid-19/issue-brief/latest-data-on-covid-19-vaccinations-by-race-ethnicity/. Published April 7, 2022. Accessed May 10, 2022.

3. Hill L, Artiga S. COVID-19 Cases and Deaths by Race/Ethnicity: Current Data and Changes Over Time. Kaiser Family Foundation. https://www.kff.org/coronavirus-covid-19/issue-brief/covid-19-cases-and-deaths-by-race-ethnicity-current-data-and-changes-over-time/. Published February 22, 2022. Accessed May 10, 2022.

4. Schöley J, Aburto JM, Kashnitsky I, et al. Bounce backs amid continued losses: Life expectancy changes since COVID-19. bioRxiv. February 2022. doi:10.1101/2022.02.23.22271380

5. de Walque Paul Gubbins Beatriz Piedad Urdinola Jeremy Veillard GDD. COVID-19 Age-Mortality Curves for 2020 Are Flatter in Developing Countries Using Both Official Death Counts and Excess Deaths. https://documents1.worldbank.org/curated/en/718461634217653573/pdf/COVID-19-Age-Mortality-Curves-for-2020-Are-Flatter-in-Developing-Countries-Using-Both-Official-Death-Counts-and-Excess-Deaths.pdf.

6. Wrigley-Field E, Kiang MV, Riley AR, et al. Geographically targeted COVID-19 vaccination is more equitable and averts more deaths than age-based thresholds alone. Sci Adv. 2021;7(40):eabj2099.

7. Situation Update for COVID-19 - Minnesota Dept. of Health - February 25, 2022. https://www.health.state.mn.us/diseases/coronavirus/situation.html. Accessed February 26, 2022.

8. Galewitz P. Health Experts Worry CDC’s Covid Vaccination Rates Appear Inflated. Kaiser Health News. <https://khn.org/news/article/cdc-senior-covid-vaccination-rates-appear-inflated/>. Published December 9, 2021. Accessed March 1, 2022.

9. Goldman N, Pebley AR, Lee K, Andrasfay T, Pratt B. Racial and ethnic differentials in COVID-19-related job exposures by occupational standing in the US. PLoS One. 2021;16(9):e0256085.

10. Faust JS, Renton B, Essien UR, Gounder CR, Lin Z, Krumholz HM. Racial and ethnic disparities in COVID-19 vaccinations in the United States during the booster rollout. bioRxiv. December 2021. doi:10.1101/2021.12.12.21267663

11. Bailey ZD, Feldman JM, Bassett MT. How Structural Racism Works - Racist Policies as a Root Cause of U.S. Racial Health Inequities. N Engl J Med. 2021;384(8):768–773.

12. McClure ES, Vasudevan P, Bailey Z, Patel S, Robinson WR. Racial Capitalism Within Public Health-How Occupational Settings Drive COVID-19 Disparities. Am J Epidemiol. 2020;189(11):1244–1253.

13. Chen YH, Glymour M, Riley A, et al. Excess mortality associated with the COVID-19 pandemic among Californians 18-65 years of age, by occupational sector and occupation: March through November 2020. PLoS One. 2021;16(6):e0252454.

14. Macias Gil R, Marcelin JR, Zuniga-Blanco B, Marquez C, Mathew T, Piggott DA. COVID-19 Pandemic: Disparate Health Impact on the Hispanic/Latinx Population in the United States. J Infect Dis. 2020;222(10):1592–1595.

15. Bile R, Gilbert A, Mohamed S, Mohammed I, Plummer M, Wrigley-Field E. Lessons From An Immigrant-Focused Community COVID-19 Vaccination Organization. Health Affairs Forefront. https://www.healthaffairs.org/do/10.1377/forefront.20220518.186581. Published May 20, 2022. Accessed June 9, 2022.

16. Centers for Disease Control and Prevention. Demographic Characteristics of People Receiving COVID-19 Vaccinations in the United States. COVID Data Tracker. https://covid.cdc.gov/covid-data-tracker/#vaccination-demographic. Accessed June 15, 2022.

17. Wrigley-Field E, Berry KM, Persad G. Race-specific, U.S. state-specific COVID-19 vaccination rates adjusted for age. medRxiv. November 2021. doi:10.1101/2021.11.19.21266612

18. Samuel H. Preston, Patrick Heuveline, Michel Guillot. Demography: Measuring and Modeling Population Processes. Wiley-Blackwell; 2000.

19. Wrigley-Field E, Garcia S, Leider JP, Robertson C, Wurtz R. Racial Disparities in COVID-19 and Excess Mortality in Minnesota. Socius. 2020;6:2378023120980918.

